# Nipah vaccine and monoclonal antibody introduction: Understanding endemic and at-risk country perspectives to inform demand forecasting

**DOI:** 10.64898/2026.09.21.26363632

**Authors:** Katelyn A. Dinkel, Anika Ruisch, Ishani Mathur, Rebakaone T. Bowe, Peter Windus, Justice Nonvignon, Damian Walker, Andrew A. Torkelson

## Abstract

**Background:** Nipah virus (NiV) is a zoonotic pathogen in the *Henipavirus* genus that causes severe, often fatal disease in humans. The World Health Organization and the Coalition for Epidemic Preparedness Innovations (CEPI) have designated NiV as a priority pathogen with pandemic potential. While CEPI is funding research on medical countermeasures, including vaccines and monoclonal antibodies (mAbs), there remains limited understanding of how at-risk countries will use them. This study aims to address this gap through targeted interviews and demand forecasting for 10 countries: Australia, Bangladesh, Cambodia, India, Indonesia, Malaysia, the Philippines, Singapore, Thailand, and Vietnam.

**Methodology/Principal Findings:** Interviews with country and global stakeholders gathered data on regional sentiment, financing, and affordability. Profiles were developed to summarize each country’s perceived outbreak risk and their likely use of countermeasures. Demand was estimated by integrating these perspectives into models of likely country-level responses to three NiV outbreak scenarios: Expected, Amplified, and Extended. In the Expected scenario, countries focused on reactive use with global annual demand ranging from approximately 800 to 5,000 doses for vaccine-only, mAb-only, and combination interventions. Ring strategies were preferred given outbreak size and infrequency. Demand was highest for vaccine-only and combination interventions, with lower demand for mAbs-only due to affordability concerns and less familiarity in Bangladesh and India. In Amplified and Extended scenarios, countries expanded their responses to include preventive campaigns for healthcare workers and rural populations. These campaigns would require approximately 410,000 and 6,300,000 doses, respectively, if introduced in all 10 countries.

**Conclusions/Significance:** This study has revealed existing sentiments regarding Nipah disease and interventions and modeled demand across three outbreak scenarios. Given global annual demand based on current epidemiology, Nipah countermeasure markets are unlikely to be commercially viable solely on routine country-financed demand. Instead, market health will require external financing and stockpile mechanisms to maintain demand between sporadic outbreaks.

**Author Summary:** Nipah virus (NiV), which causes severe and often fatal disease in humans, is recognized by the Coalition for Epidemic Preparedness Innovations (CEPI) as a priority pathogen with pandemic potential. In response, CEPI has supported the development of vaccines and monoclonal antibodies (mAb). However, limited evidence exists on how at-risk countries will deploy these medical countermeasures. We interviewed key decision-makers across 10 at-risk countries, along with global experts on NiV, to better understand disease prioritization, financing and affordability considerations, and countermeasure preferences. Interview findings informed the development of a demand forecast in response to Nipah outbreaks of varying intensity. At expected outbreak levels, countries focused their response on reactive strategies given outbreak size and infrequency. As outbreaks grew in size, countries considered expanding their response to include preventive campaigns targeting healthcare workers and at-risk, rural populations. This study provides new insights into country-level perspectives on NiV and medical countermeasures and quantifies expected demand across three outbreak scenarios and 10 countries. Our findings can provide a foundation for the strategic development of an international Nipah countermeasure stockpile, designed to address episodic NiV outbreaks and enhance pandemic preparedness.

## 1. Introduction

Nipah virus (NiV) is a zoonotic pathogen in the *Henipavirus* genus (*Paramyxoviridae* family) that causes severe, often fatal, illness in humans. First identified during a 1998–1999 outbreak among pig farming communities in Malaysia and Singapore, it caused acute encephalitic and respiratory illness, resulting in over 100 human fatalities and the mass culling of more than one million pigs [1]. Fruit bats in the Pteropus genus were subsequently identified as the principal reservoir [2,3]. These bats are widely distributed across South and Southeast Asia and shed the virus in saliva and urine, creating repeated opportunities for zoonotic spillover through contaminated food sources and shared environments.

Since 2001, NiV outbreaks have predominantly occurred in Bangladesh and India [3], with transmission driven largely by direct bat-to-human contact and secondary human-to-human spread. Outbreaks have been characterized by low transmissibility (effective reproduction numbers average 0.33) and high case fatality rates (CFR; estimates ranging from 40% to 90%) [4,5]. Despite low transmissibility, there has been repeated evidence of nosocomial and household transmission [4,6], with sustained interpersonal transmission in healthcare and community settings [7,8].

NiV has been designated a priority pathogen with pandemic potential by the World Health Organization (WHO) and the Coalition for Epidemic Preparedness Innovations (CEPI) [9,10]. In response, CEPI has supported the development of medical countermeasures (MCMs) as part of global epidemic preparedness efforts. Currently, no approved vaccines or monoclonal antibody (mAb) therapies are available, but several candidates are in clinical development, including the University of Oxford’s Phase 2 vaccine, Public Health Vaccines’ Phase 2 vaccine, and Mapp Biopharmaceutical’s Phase 1 single-dose mAb for prophylaxis and treatment [11,12].

Although these candidates mark significant scientific progress, their effectiveness will ultimately depend on practical deployment, particularly in low- and middle-income countries (LMICs) where NiV risk is highest. NiV’s outbreak characteristics, combining low transmissibility with a high CFR, uniquely position it among emerging pathogens and necessitate tailored strategies for MCM deployment. Drawing on lessons from previous outbreak responses is critical to shaping effective strategies for NiV prevention and control. Experiences from the management of Ebola and COVID-19 have demonstrated that timely, evidence-based reactive and proactive approaches can significantly reduce transmission and mortality [13–15]. These lessons are applicable to NiV, where targeted ring strategies and preventive campaigns for high-risk groups (e.g., HCWs or rural communities near known spillover zones) could enhance preparedness to contain outbreaks before they escalate.

Despite increasing recognition of NiV as a priority pathogen of epidemic and pandemic potential, limited understanding exists regarding how vaccines and mAbs could be used in LMICs and endemic regions. Although several vaccine and mAb candidates are advancing through early clinical development with support from global partnerships, the path from development to access requires attention. Data on anticipated demand, regional sentiment, financing mechanisms, familiarity with mAbs, and affordability constraints are scarce, yet these considerations are essential for shaping evidence-based strategies for product deployment, regulatory preparedness, manufacturing scale-up, equitable access, and stockpiling.

This study aims to address these questions by analyzing country- and regional-level sentiment, financial factors, affordability issues, and demand for potential NiV vaccines and mAbs from the perspectives of select countries in the endemic region. It focuses on Australia, Bangladesh, Cambodia, India, Indonesia, Malaysia, the Philippines, Singapore, Thailand, and Vietnam. The results provide an initial demand forecast to guide strategic planning for equitable vaccine and mAb access, and to inform future policy and investment decisions supporting global preparedness for Nipah and related emerging pathogens.

## 2. Methods

This study used a mixed-methods approach to assess demand, country- and global-level sentiment, financing, and affordability for Nipah MCMs from the perspectives of LMICs and endemic regions. The objective was to generate early insights into strategic considerations for the deployment of vaccines and mAbs for Nipah, supporting CEPI’s planning for global, equitable access to future countermeasures.

The study followed a stepwise approach, combining (1) desk review, (2) key informant interviews (KIIs) with country and global stakeholders, and (3) demand modeling, with each step based on the results from previous steps. Findings from the desk review informed the interview guides. Results from the desk review and KIIs were synthesized into country- and global-level assessments, which were subsequently used to parameterize a demand forecast model under multiple outbreak scenarios.

### 2.1. Country Selection

10 countries in the Asia-Pacific region were included to represent the following categories: (1) settings with active or recurrent outbreaks (Bangladesh, India); (2) settings with documented past outbreaks (Malaysia, the Philippines, Singapore); and (3) settings with ecological or epidemiological risk for future outbreaks but no known outbreaks to date (Australia, Cambodia, Indonesia, Thailand, Vietnam). All at-risk countries were not included in the study, with selection contingent on the feasibility of stakeholder engagement.

### 2.2. Country- and Global-Level Assessments

The country-level and global-level assessments applied a combination of desk research and KIIs to assess NiV preparedness, response capacity, and decision-making processes and develop profiles for each country in this analysis. The assessments were guided by the WHO framework for decision-making on new vaccine introduction, which considers: (1) disease burden and public health priority; (2) characteristics and constraints of available products; and (3) health system capacity [16].

#### Desk Research

A structured review compiled information on NiV epidemiology, surveillance and laboratory systems, immunization platforms, regulatory pathways, financing mechanisms, and health system capacity. Sources included peer-reviewed literature, government documents, global databases, grey literature, and media reports. Searches combined disease-specific terms (“Nipah virus,” “henipavirus”) with preparedness-related terms (“surveillance,” “vaccine,” “monoclonal antibody,” “financing,” “stockpiling”) and country names.

#### Key Informant Interviews (KIIs)

Semi-structured interview guides were developed based on desk findings and WHO guidance. Seven domains were explored: public health and political priorities; disease burden and awareness; prevention and response measures; decision-making structures; financing and affordability; supply chain and stockpiling; and contextual considerations. Virtual interviews were conducted in English between June–September 2025 (country-level) and May–November 2025 (global-level), each lasting 30–60 minutes. Informed consent was obtained from all participants. Individual names have been withheld to maintain confidentiality. One respondent completed an online survey instead of an interview.

Country stakeholders were purposively sampled to include ministry officials, regulators, National Immunization Technical Advisory Group (NITAG) members, and research institutions. A total of 14 KIIs and one online survey were completed across seven countries. Interviews were conducted in Bangladesh (one), India (four), Indonesia (two), Malaysia (two, including one group interview with six participants), the Philippines (two interviews plus one survey), Singapore (two), and Thailand (one). For Australia, Cambodia, and Vietnam, country profiles were developed solely through desk research after repeated outreach to potential stakeholders did not result in confirmed interview participation during the study period. In addition, 16 global stakeholders from international organizations like Gavi and the WHO, nine health agencies, and academic institutions were interviewed to inform perspectives on prioritization, outbreak risk, and deployment considerations. The interview guides used in the country- and global- assessments are included in the Supporting Information (S1 and S2, respectively).

#### Ethics Statement

Management Sciences for Health’s Scientific Committee assessed the proposed protocol to verify whether the planned activity meets the definition of Human Subjects Research and therefore requires review by an Institutional Review Board (IRB). The Scientific Committee determined that the Nipah stakeholder interviews did not meet the definition of Human Subjects Research and therefore did not require submission to an IRB. This was decided because the study was designed to collect information on national policies on pandemic preparedness and funding pathways for new vaccines; as such, it did not ask participants for information about themselves. Ethical principles and appropriate protections were maintained throughout the data collection, storage, assessment, and analysis process.

### 2.3. Demand Forecast

Using data collected from the country profiles and stakeholder inputs, a deterministic demand model was developed to estimate the number of vaccine and mAb doses required under different outbreak scenarios.

#### Intervention assumptions

Since Nipah interventions are in the early stages of development, demand scenarios for vaccines and mAbs were developed in line with the WHO’s target product profile and incorporated country preferences from KIIs. Both vaccines and mAbs were assumed to follow a single-dose injectable course, elicit protection within 2 weeks, and provide protection for at least 6 months and ideally for more than 1 year [17]. Vaccines were assumed to be used for immunizing at-risk groups at any age. mAbs were assumed to be used for pre- and post-exposure prophylaxis and for therapeutic purposes in all cases. Three intervention configurations were modeled for each outbreak scenario, including vaccines only, mAbs only, and a combination of vaccines and mAbs.

#### Scenarios

Three demand scenarios were developed to evaluate each target county’s expected response to outbreaks of varying sizes (Table 1). Country- and state-level burden data were extracted from published literature, and Monte Carlo simulations were used to estimate expected annual case counts and outbreak size distributions. The Expected outbreak (Scenario 1) estimated demand based on current epidemiology in countries with historically recurrent outbreaks. Current epidemiology estimates from a country-level Monte Carlo simulation included annual recurrent cases in Bangladesh (7), India (2), and Malaysia (2). Zero cases were assumed for all other countries in the analysis. In this scenario, demand was aggregated across all outbreak countries to estimate global annual demand, assuming the outbreaks occurred in the same year.

**Table 1.**
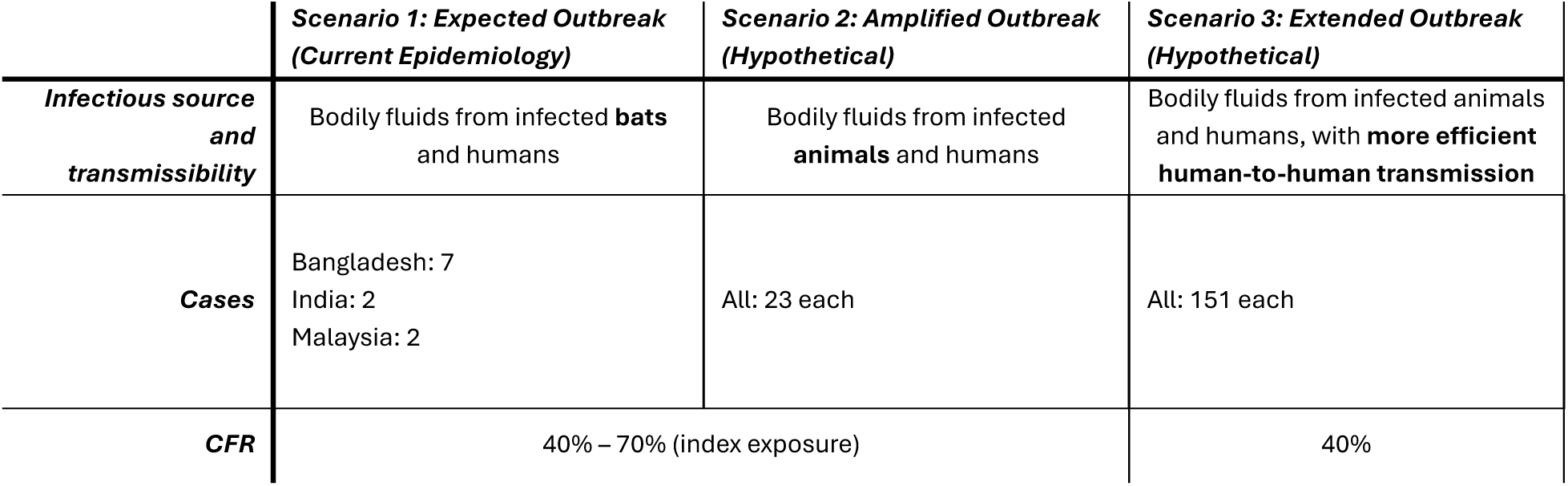
Nipah outbreak scenarios with infectious source, transmissibility, and CFR linked to outbreak. The Expected outbreak (Scenario 1) examined the response to expected annual outbreaks given current global case burden. The Amplified and Extended outbreaks (Scenarios 2-3) examined the modeled demand from a hypothetical Nipah outbreak in each country in the analysis.

The Amplified (Scenario 2) and Extended (Scenario 3) outbreaks were largely consistent with scenario definitions from a concurrent study of NiV vaccine and mAb impact [18]. Scenario assumptions across Amplified and Extended outbreaks differed by source, transmissibility, outbreak size, and CFR. Outbreak size assumptions were derived from a combined Monte Carlo simulation that aggregated all reported past outbreaks across countries into a single dataset. Case counts were summed across countries within a calendar year and then used to generate a distribution of case counts for larger outbreaks. Results from the Monte Carlo simulation of this dataset yielded 23 cases for an Amplified outbreak (the median outbreak) and 151 cases for an Extended outbreak (the 99th-percentile outbreak). In these scenarios, results were grouped into three categories: active outbreaks (Bangladesh and India), past outbreaks (Malaysia, Philippines, and Singapore), and no known outbreaks (Australia, Cambodia, Indonesia, Thailand, and Vietnam). Demand was reported at the country level because each country’s response was modeled for an isolated, hypothetical outbreak within its borders. These outbreaks were not assumed to occur simultaneously across countries, nor were outbreaks in one country assumed to influence responses in another. Further details on the burden data and Monte Carlo simulations are provided in the Supporting Information (S3).

#### Country Response Assumptions, Target Population, and Demand Calculation

Individual country response strategies were informed by country profiles, with assumptions on response type, intervention intensity, coverage, and scope tailored to KII findings. Countries with stronger preparedness and higher income were modeled with higher coverage levels [19,20].

Target populations included in the analysis were Nipah patients (case therapeutics), ring strategies, HCWs, and rural populations (Table 2). Reactive responses included case therapeutics and ring strategies targeting contacts and contacts-of-contacts. Ring strategies were estimated using the median number of contacts (17) from a contact tracing study in Bangladesh [5]. Proactive responses included preventive campaigns to reach HCWs and rural, male populations at risk of spillover. HCWs were included due to their elevated risk from closer encounters with infected patients in hospital environments [21]. Rural male populations were targeted because men accounted for 74% of human cases with available demographic data in Sun et al. [22]. Full details on the target population estimates are provided in the Supporting Information (S4). Full coverage and buffer assumptions for each country, scenario, and target population are provided in the Supporting Information (S5).

**Table 2.** Demand forecasting assumptions by target population.

| **TARGET POPULATION** | **TARGET POPULATION DATA SOURCE** | **COVERAGE ASSUMPTION** | **COVERAGE SOURCE** | **BUFFER ASSUMPTION** | **BUFFER SOURCE** |
| --- | --- | --- | --- | --- | --- |
| **NIPAH CASES** | Monte Carlo estimates | 80-100% | Katz et al. 2011 [23] | 50% | Estimate; assume higher order for uncertain amount of cases [24] |
| **RING STRATEGY: IMMEDIATE CONTACTS AND CONTACTS OF CONTACTS** | Median number of contacts (17) from Nikolay et al. 2019 [5] | 80-100% | Perera, et al. 2024 [25] | 50% | Estimate; assume higher order for uncertain amount of cases [24] |
| **HCWS** | WHO Health Workforce statistics database [26]; Health Labour Market Analysis in Bangladesh [27] | 30-60% | Bisanzio, et al. 2023 [28] | 25% | WHO Cold Chain and Logistics Management Handbook [29] |
| **RURAL POPULATIONS** | UN World Urbanization Prospects [30]; UN World Urbanization Prospects [31]; Assumes male population is 50% of total population | 5-10% | Bisanzio, et al. 2023 [28] | 25% | WHO Cold Chain and Logistics Management Handbook [29] |

For each country *i*, the total demand in response to a specific outbreak *j* was calculated as:

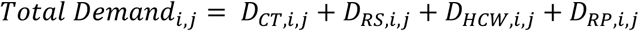

Where:

- *Total Demand_i,j_* = Total demand across all demand sources for country *i* in response to outbreak *j*
- *D_CT,i,j_* = Demand for case therapeutics for country *i* in response to outbreak *j*
- *D_RS,i,j_* = Demand for ring strategies for country *i* in response to outbreak *j*
- *D_HCW,i,j_* = Demand for HCWs for country *i* in response to outbreak *j*
- *D_RP,i,j_* = Demand for male rural populations for country *i* in response to outbreak *j*

The demand estimates for each target population in country *i* was calculated as:

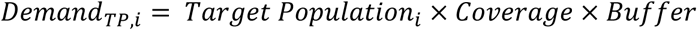

Where:

- *Demand_TP,i_* = number of vaccines or mAbs needed for target population in country *i*
- *Target Population_i,_* = size of specific population eligible for response strategy in country *i*
- *Coverage* = proportion of the target population receiving intervention in country *i*
- *Buffer* = additional proportion to account for wastage and uncertainty in country *i*

Demand in each scenario was estimated at the subnational level, focusing on states at the first administrative level considered most at risk for human NiV infection. In India and Bangladesh, high-risk states were identified as those with expected annual cases from the country/state Monte Carlo simulation. In Cambodia, Indonesia, Malaysia, the Philippines, Singapore, and Thailand, states were selected based on historical human cases or seropositive animal reservoirs [22]. For Vietnam and Australia, where seropositivity data were limited, state selection was guided by the presence of Pteropus bat reservoirs [32]. Population estimates and selected subnational areas are detailed in the Supporting Information (S6).

Demand estimates for each target population were rounded to two significant figures. Final aggregate numbers, including global-level estimates for the Expected scenario and country-level estimates for the Amplified and Extended scenarios, were calculated by summing demand across all applicable target populations and then rounding by magnitude (nearest 10 for values up to 1,000, nearest 100 for values between 1,000 and 10,000, nearest 1,000 for values between 10,000 and 100,000, nearest 10,000 for values between 100,000 and 1,000,000, and nearest 100,000 for values greater than or equal to 1,000,000). Demand values for the Expected scenario were summed across all countries to estimate global demand for current epidemiology. Demand values for the Amplified and Extended scenarios were summed at the country-level since outbreaks were assumed to be isolated events within each country. A sensitivity analysis was conducted by varying the assumptions for coverage and buffer for each target population and outbreak scenario in each country. Demand values were directly proportional to these assumptions. The full results of this sensitivity analysis for response type by country are included in the Supplementary Information (S7).

The demand models were built in Microsoft Excel software (Microsoft Corp., Redmond, Washington, US).

## 3. Results

### 3.1. Key Informant Interview Results

#### Country Findings

Country findings varied based on outbreak history, perceived risk, and health system capacity, but several patterns emerged across settings with active, past, or no known outbreaks to date. In general, outbreak experience was associated with higher awareness and clearer emergency financing pathways. Countries without outbreak experience typically integrated Nipah into broader zoonotic surveillance systems without dedicated preparedness measures. These findings are summarized in Table 3, and detailed country profiles are available in the Supporting Information (S8).

**Table 3.** Summary of country findings. Institutional acronyms are defined in a note below the table.

| Country | Income Level | Total expenditure on vax financed by govt [33] | Outbreak Status | Surveillance & Preparedness | Preferred MCM (Vaccines vs. mAbs) | Financing & Emergency Funding* | Stockpiling & Access | Key Takeaways |
| --- | --- | --- | --- | --- | --- | --- | --- | --- |
| **Bangladesh** | LMI | 49% | Active outbreaks | Hospital-based surveillance (CDC/IEDCR-supported); limited laboratory capacity | No clear overall preference; mAbs considered useful for small caseloads | IDA-eligible; heavy donor reliance; affordability major concern | Minimal stockpiling; unclear disease prioritization | Regular small outbreaks; donor dependence; affordability barrier |
| **India** | LMI | 99% | Active outbreaks | Surveillance led by The Integrated Disease Surveillance Programme; strong diagnostics; state-level variation | Vaccines preferred (mAbs considered costly and limited in utility for rapid progression) | ADB/APDRF eligible; emergency funds available | Emerging manufacturing capacity; no Nipah-specific stockpile | Large domestic capacity; dual state-central fragmentation |
| **Malaysia** | UMI | No Data | Past outbreak (1998) | Strong One Health system; notifiable disease; high institutional awareness | No explicit preference; mAbs noted for curative appeal | Emergency funds via MoH/MoF; ASEAN access | Existing national stockpile system (not Nipah-specific) | Strong legacy capacity and laboratory capacity for Nipah diagnostics exists |
| **Philippines** | LMI | 100% | Past outbreak (2014) | Zoonotic surveillance; low current prioritization | No stated preference; concerns around costs of mAbs | Financing subject to Health Technology Assessment review and price negotiation; ASEAN/ADB/APDRF eligible | No vaccine or therapeutic stockpile; PPE and diagnostics only | Regulatory reform ongoing; donor dependence; affordability barrier |
| **Singapore** | HI | No Data | Past outbreak (1990s) | Disease-agnostic pandemic framework; rapid reporting; strong clinical capacity | Combination preferred | High fiscal flexibility; national reserves accessible; ASEAN member | Dynamic stockpile model including essential MCMs | Highly organized, adaptable system; outbreak readiness (not Nipah specific) |
| **Australia** | HI | 100% | No known outbreak | Robust laboratory and zoonotic surveillance | No response** | Emergency funds under EADRA; not IDA-eligible | National medical stockpile (details not public) | High readiness, excellent capacity, low perceived risk |
| Indonesia | UMI | 90% | No known outbreak | Integrated One Health surveillance; limited Nipah-specific lab capacity | Vaccines preferred (mAbs costly, logistically challenging) | Emergency funds accessible upon outbreak declaration; ASEAN/ADB/APDRF eligible | No Nipah-specific stockpile; cold chain constraints noted | Donor dependence; affordability barrier |
| Thailand | UMI | 100% | No known outbreak | Strong bat surveillance under One Health; limited policy prioritization | No explicit preference | ASEAN/APDRF eligible; no Nipah-specific allocation | No Nipah-specific stockpile | Awareness present but low readiness; potential risk via bats |
| Vietnam | LMI | 67% | No known outbreak | Bat circulation documented; not a priority zoonosis | No response** | IDA transitional; ASEAN/ADB/APDRF eligible | No Nipah-specific stockpile | Strong domestic vaccine base; potential risk via bats |
| Cambodia | LMI | 38% | No known outbreak | Nipah included in zoonotic strategies; limited operational capacity | No response** | IDA-eligible; donor-supported preparedness | No dedicated stockpile; no regulatory pathway identified | Very low awareness; donor dependence; affordability barrier |

\*List of institutional acronyms: U.S. Centers for Disease Control and Prevention (CDC); Institute of Epidemiology, Disease Control and Research (IEDCR); International Development Association (IDA); The Association of Southeast Asian Nations (ASEAN); Asian Development Bank (ADB); Emergency Animal Disease Response Agreement (EADRA); Asia Pacific Disaster Response Fund (APDRF)
\*\*No response: No KIIs were conducted. Information on other topics for these countries was collected from a desk review.

In countries experiencing active outbreaks (Bangladesh and India), Nipah was recognized as a recurring public health threat, though responses varied in scale and financing capacity. The Bangladeshi interviewee reported that annual cases exhibited high CFRs and that the hospital-based surveillance system was supported by external partners. However, domestic financing for vaccines, stockpiles, or other MCMs remained uncertain, with costs a key barrier. In India, interviewees also reported recurring cases in specific states, with surveillance integrated into district hospital systems. They reported that the government has the capacity to mobilize emergency funds and covers a substantial share of vaccine procurement costs. Nevertheless, Nipah remained a lower priority compared to other zoonoses due to the limited number of cases. Stakeholders in both countries generally favored vaccines over mAbs, citing the perceived high cost of mAbs and limited domestic manufacturing, while acknowledging that mAbs may be suitable in low-transmission settings. Bangladeshi officials indicated that budget constraints were a major concern for introducing mAbs.

Among countries with past outbreaks, including Malaysia, the Philippines, and Singapore, prior experience influenced awareness, though current priorities varied. Malaysia’s 1998 outbreak resulted in sustained institutional awareness and notifiable disease status [27], but Nipah was not a current government focus. Interviewees explained that emergency funding mechanisms exist through national ministries and regional platforms. They also noted the therapeutic potential of mAbs but did not express a clear preference for them over vaccines. In the Philippines, officials indicated that a single outbreak led to limited preparedness, with no dedicated stockpiling or prevention measures despite functional baseline infectious disease systems. Future use of MCMs would depend on health technology assessments, emergency flexibilities, and evidence of burden or cost-effectiveness, without specific product preferences. In Singapore, despite no recent cases, high preparedness persisted under a disease-agnostic pandemic framework supported by strong surveillance and laboratory systems. Participants indicated a preference for a context-dependent approach, using mAbs for small outbreaks and vaccines for larger-scale events, with emergency funding and flexible stockpiling mechanisms readily available, including regional engagement through the Association of Southeast Asian Nations (ASEAN) platforms.

In countries with no known outbreaks to date, including Australia, Cambodia, Indonesia, Thailand, and Vietnam, Nipah was generally monitored within broader zoonotic or One Health frameworks. Nonetheless, it has not been a top public health priority for these countries. Australia classified Nipah as a notifiable animal disease and maintained emergency funding mechanisms for zoonotic threats [34]. Cambodia included Nipah as a priority in its national zoonotic strategies [35]. In Indonesia, the interviewee explained that preparedness was limited and that deployment of MCMs would depend on a formal outbreak declaration to activate emergency funding. The interviewee also noted that, in general, vaccines were considered more feasible than mAbs given Indonesia’s archipelagic geography. Thailand maintains strong laboratory and manufacturing capacity, along with long-standing bat surveillance, but the interviewee did not disclose a clear preference between vaccines and mAbs. Vietnam has adopted a precautionary preparedness posture, classifying NiV as a Group A notifiable disease that requires immediate reporting and infection prevention and control (IPC) measures. Surveillance has been enhanced at points of entry and within healthcare facilities, with NiV detection integrated into platforms monitoring encephalitis and severe respiratory illnesses. Referral pathways to Biosafety Level-3 labs have been established [36].

#### Global/Regional Level Findings

Global and regional stakeholder KIIs focused on perceived drivers of demand for NiV medical MCMs, as well as structural and contextual constraints that could limit uptake. Stakeholders consistently identified disease severity as a primary driver of demand. The high CFR and the risk of severe neurological sequelae were described as strong justifications for both preventive vaccines and therapeutic mAbs, even in the context of relatively small outbreaks. Zoonotic spillover risk was also emphasized. Interviewees pointed to ecological disruption, deforestation, and increased human-bat interactions as factors contributing to continued spillover risk. This sustained underlying risk was viewed as supporting ongoing preparedness investments rather than purely reactive responses. Within this risk, occupational exposure emerged as a clear priority area. HCWs, outbreak responders, and laboratory personnel were widely recognized as high-risk groups due to predictable, repeated exposure during outbreaks. Targeted vaccination or prophylactic mAb use in these populations was considered operationally feasible and justifiable, and some stakeholders suggested that limited pre-emptive immunization of HCWs and, in some contexts, other high-risk communities could form part of preparedness planning even in the absence of frequent outbreaks.

Regarding outbreak response needs, ring vaccination strategies and reactive mAb deployment were broadly supported. Stakeholders described these approaches as efficient and practical in settings with strong surveillance, case identification, and contact tracing systems. Reactive use was viewed as particularly relevant given the typically localized nature of outbreaks. Finally, pandemic potential was referenced as an important contextual factor. Recognition of NiV, and particularly the hypothetical emergence of a more transmissible variant, was cited as supporting arguments for stockpiling at both regional and global levels.

At the same time, stakeholders underscored several health system and contextual challenges that shape preparedness and response for Nipah outbreaks. The low incidence and sporadic nature of outbreaks were widely viewed as creating uncertainty for planning and sustaining investments, especially for routine preventive vaccination beyond defined high-risk groups. Infrequent and geographically concentrated outbreaks were seen as challenging for maintaining political and financial momentum. Competing public health priorities were frequently mentioned. Compared to higher-burden or more transmissible pathogens, NiV was described as ranking lower in national and global prioritization processes, particularly in resource-constrained settings.

Gaps in surveillance and diagnostics were also raised. In some regions, fragmented or under-resourced surveillance systems limit the ability to detect and respond to outbreaks quickly, affecting the feasibility and timeliness of targeted interventions such as ring vaccination or therapeutic deployment. Stakeholders further cited regulatory and delivery capacity constraints. Limited experience with complex biologics, including mAbs, and operational constraints within health systems were described as potential barriers to timely deployment and effective use during outbreak response, rather than routine introduction.

Financing and affordability emerged as major concerns. Uncertainty around product pricing, procurement pathways, and long-term financing mechanisms was viewed as a barrier to equitable access, particularly for vaccines without broader cross-protection benefits. Finally, evidence gaps were noted. Limited data on the duration of protection, cost-effectiveness, and clearly defined priority populations, which complicate policy decision-making and preparedness planning.

Together, these drivers and constraints illustrate the tension between the high perceived severity and outbreak risk of NiV and the practical challenges of sustaining preparedness for a rare, episodic disease. Approaches, therefore, must be adaptable, targeted, and responsive to uncertain and changing epidemiological patterns.

### 3.2. Demand Forecast

The demand forecast combined assumptions from KIIs with subnational target population estimates to project the potential need for NiV MCMs. Responses were modeled across three outbreak scenarios: Expected, Amplified, and Extended.

#### Scenario Responses

Country-specific KIIs informed assumptions regarding intervention preferences (vaccines vs. mAbs), disease prioritization, and response intensity (case therapeutics, reactive ring strategies, and proactive campaigns targeting HCWs and rural populations). For countries without KIIs, assumptions were inferred from similar countries based on disease burden and income status. In the Expected outbreak (Scenario 1), countries with cases (Bangladesh, India, and Malaysia) generally favored reactive strategies due to the low number of cases. For all other countries without recurrent Nipah cases, no outbreak response was assumed until the larger, hypothetical outbreaks. As outbreaks grew larger in the Amplified and Extended scenarios, country responses became more aggressive due to the perceived threat and likely global support for outbreak control. Most countries included case therapeutics and ring strategies as a baseline, with the most aggressive responses adding HCWs in an Amplified outbreak and rural populations in an Extended outbreak (e.g., India, Malaysia, Singapore, Australia, and Indonesia). More conservative and/or cost-conscious countries waited until the Extended scenario to expand their response beyond ring strategies to include HCWs (e.g., Bangladesh, Cambodia, the Philippines, Thailand, and Vietnam). All assumptions from the stakeholder interviews are shown in Table 5, broken down by country group, country, intervention, and scenario.

**Table 4.**
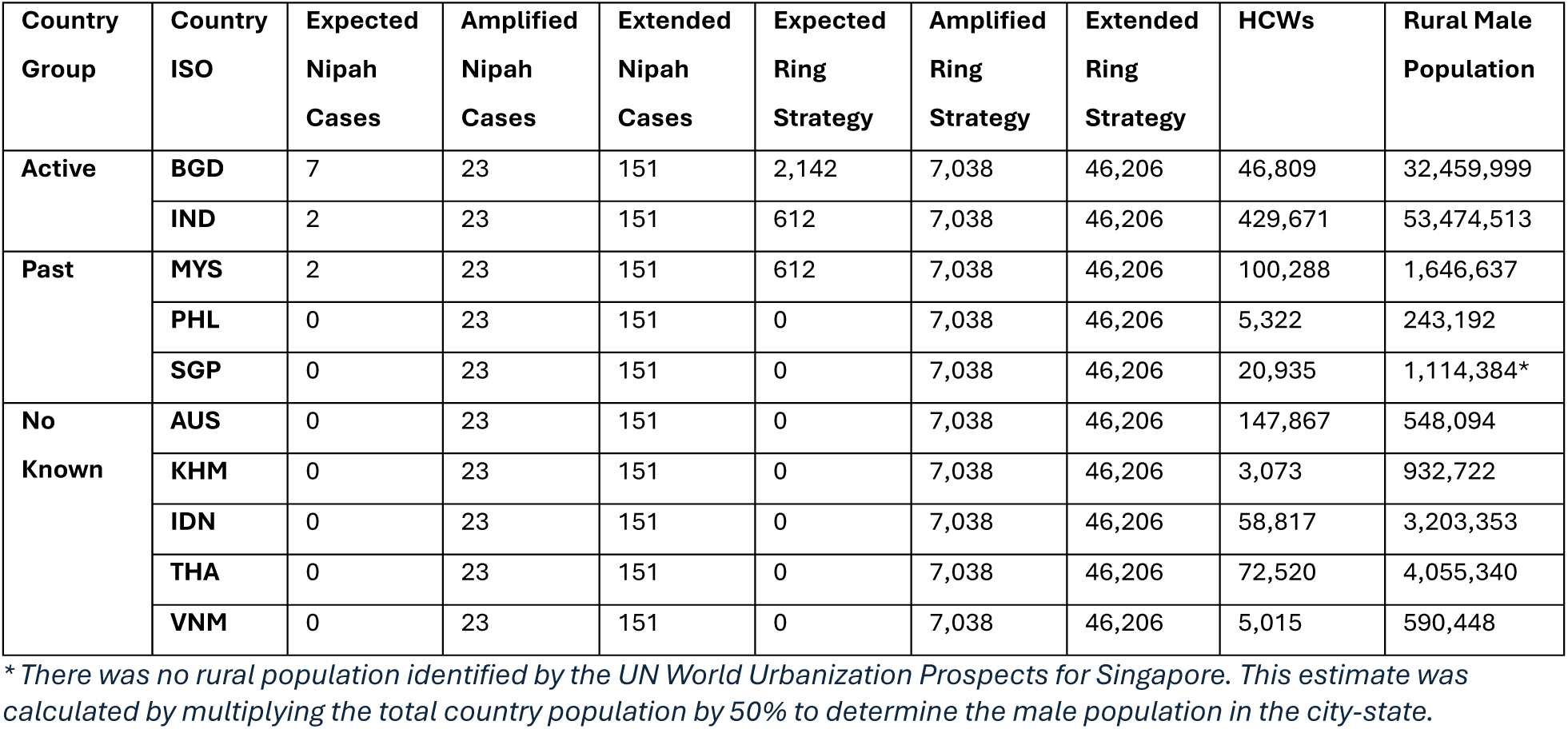
Target populations of interest in NiV outbreak response strategies, organized by country group and country.

| Country Group | Country ISO | Expected Nipah Cases | Amplified Nipah Cases | Extended Nipah Cases | Expected Ring Strategy | Amplified Ring Strategy | Extended Ring Strategy | HCWs | Rural Male Population |
| --- | --- | --- | --- | --- | --- | --- | --- | --- | --- |
| Active | BGD | 7 | 23 | 151 | 2,142 | 7,038 | 46,206 | 46,809 | 32,459,999 |
|  | IND | 2 | 23 | 151 | 612 | 7,038 | 46,206 | 429,671 | 53,474,513 |
| Past | MYS | 2 | 23 | 151 | 612 | 7,038 | 46,206 | 100,288 | 1,646,637 |
|  | PHL | 0 | 23 | 151 | 0 | 7,038 | 46,206 | 5,322 | 243,192 |
|  | SGP | 0 | 23 | 151 | 0 | 7,038 | 46,206 | 20,935 | 1,114,384* |
| No Known | AUS | 0 | 23 | 151 | 0 | 7,038 | 46,206 | 147,867 | 548,094 |
|  | KHM | 0 | 23 | 151 | 0 | 7,038 | 46,206 | 3,073 | 932,722 |
|  | IDN | 0 | 23 | 151 | 0 | 7,038 | 46,206 | 58,817 | 3,203,353 |
|  | THA | 0 | 23 | 151 | 0 | 7,038 | 46,206 | 72,520 | 4,055,340 |
|  | VNM | 0 | 23 | 151 | 0 | 7,038 | 46,206 | 5,015 | 590,448 |

\* There was no rural population identified by the UN World Urbanization Prospects for Singapore. This estimate was calculated by multiplying the total country population by 50% to determine the male population in the city-state.

**Table 5.**
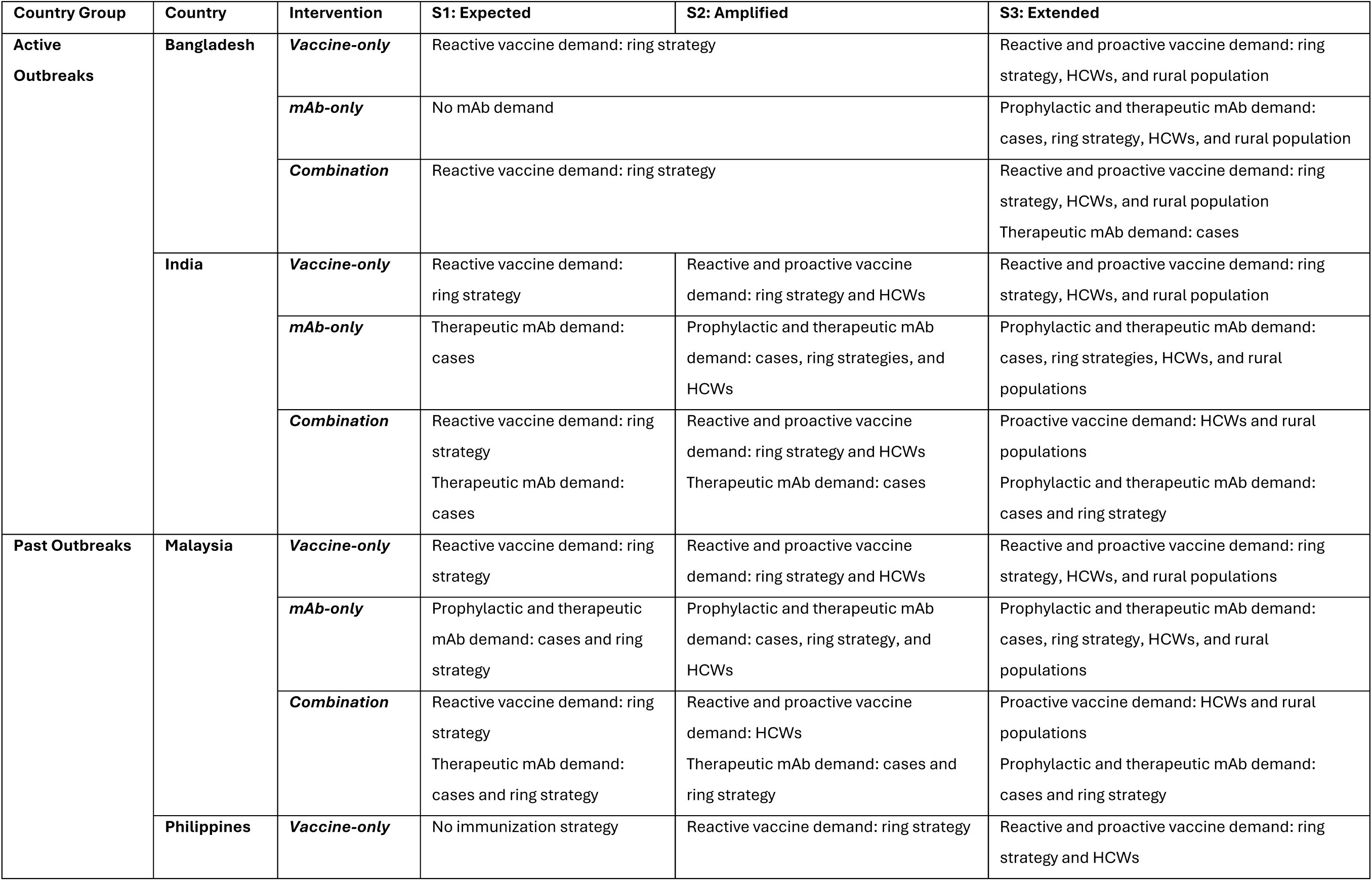
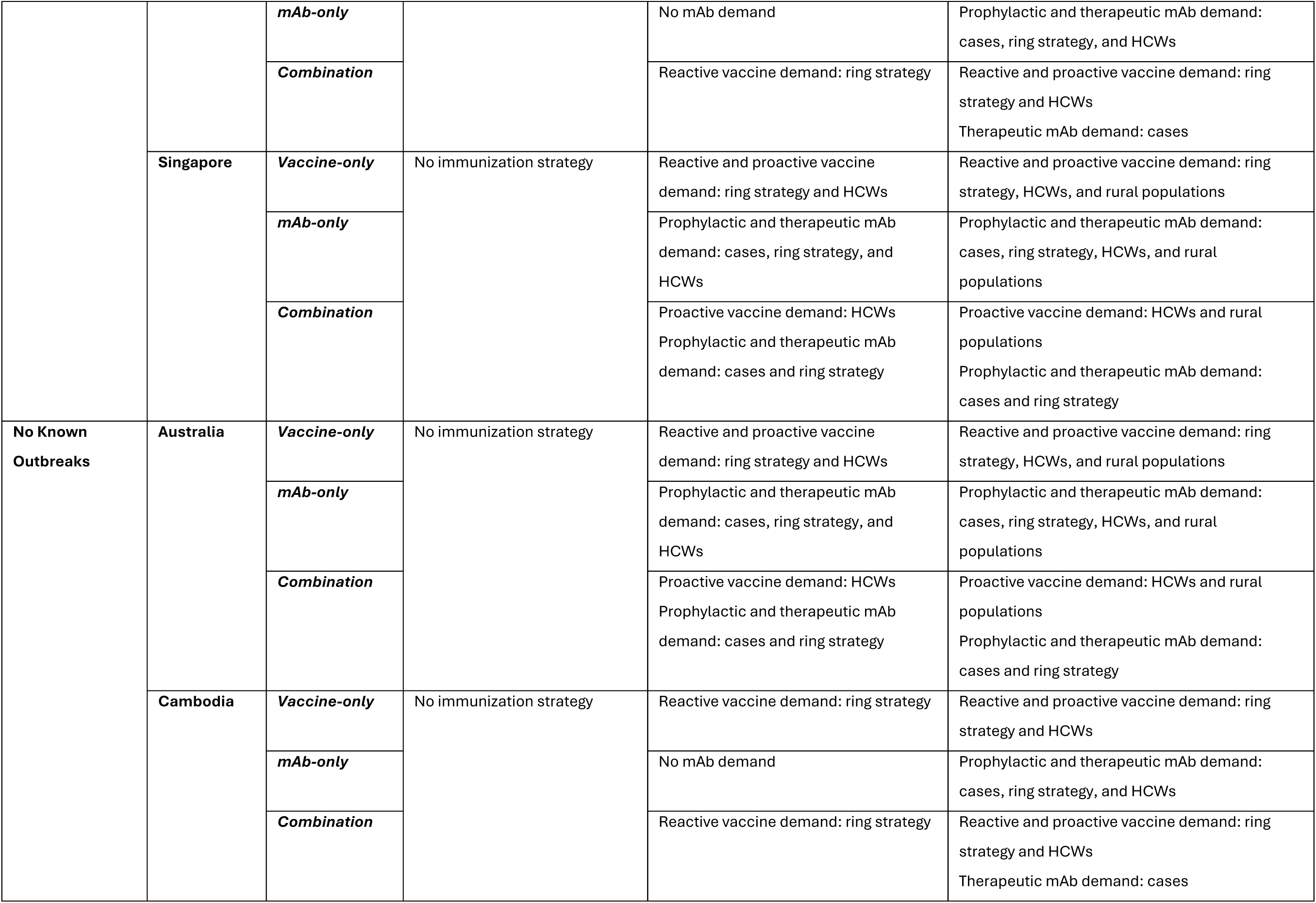
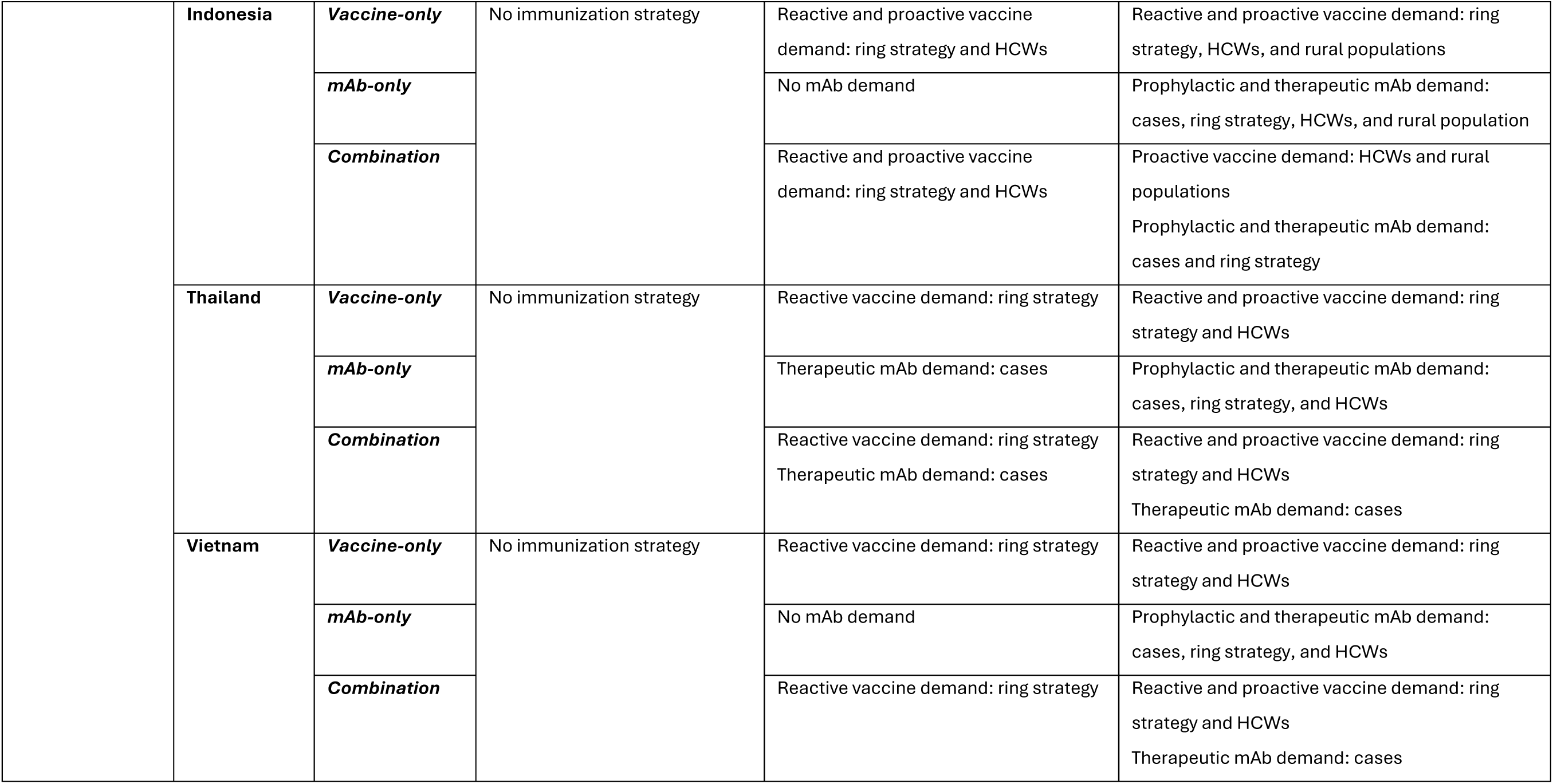
Response assumptions from country-stakeholder KIIs, organized by country group, country, intervention, and scenario.

#### Target Population Estimates

Target population estimates were generated for each country of interest, including calculations for Nipah cases, ring strategies, HCWs, and rural male populations. Target populations for ring strategies were determined by the number of cases and the assumed number of contacts per person (17), resulting in similar estimates across countries. Target populations for HCWs and rural populations depended on the size of the country’s health system, overall population, and geographic spread of cases. The active outbreak countries (Bangladesh and India) had the highest combined target population for proactive approaches among all groups, reflecting their centrality to any Nipah MCM market. The complete list of target populations by country group and individual country is provided in Table 4.

#### Demand Estimates

##### Global Annual Demand for Expected Outbreaks (Scenario 1)

Based on expected annual outbreaks limited to two Indian states (Kerala and West Bengal), four Bangladeshi districts (Dhaka, Khulna, Mymensingh, and Rangpur), and Malaysia, global annual demand ranged from 830 to 4,600 doses (Figure 1). Response in the three affected countries focused on a reactive approach, given the number of cases and the infrequency of outbreaks. The highest demand occurred when available interventions included only vaccines or a combination of vaccines and mAbs (4,600 doses). The vaccine-only intervention used only vaccines for a ring strategy, while the combination intervention paired a vaccine ring strategy with mAbs for case therapeutics. Vaccines accounted for 99% of total demand in this combination intervention response. In the mAb-only intervention, demand was lower (830 doses) due to limited use in Bangladesh and India, driven by affordability and familiarity concerns. In vaccine-only and combination interventions, Bangladesh had the highest vaccine demand due to a higher expected caseload (7), requiring 2,900 doses for a vaccine ring strategy. India and Malaysia anticipated two cases each, needing approximately 1,700 doses combined to cover their ring strategies. Malaysia was the only country to favor strong mAb use, including mAbs in a ring strategy in the mAb-only intervention scenario.

**Figure 1.**
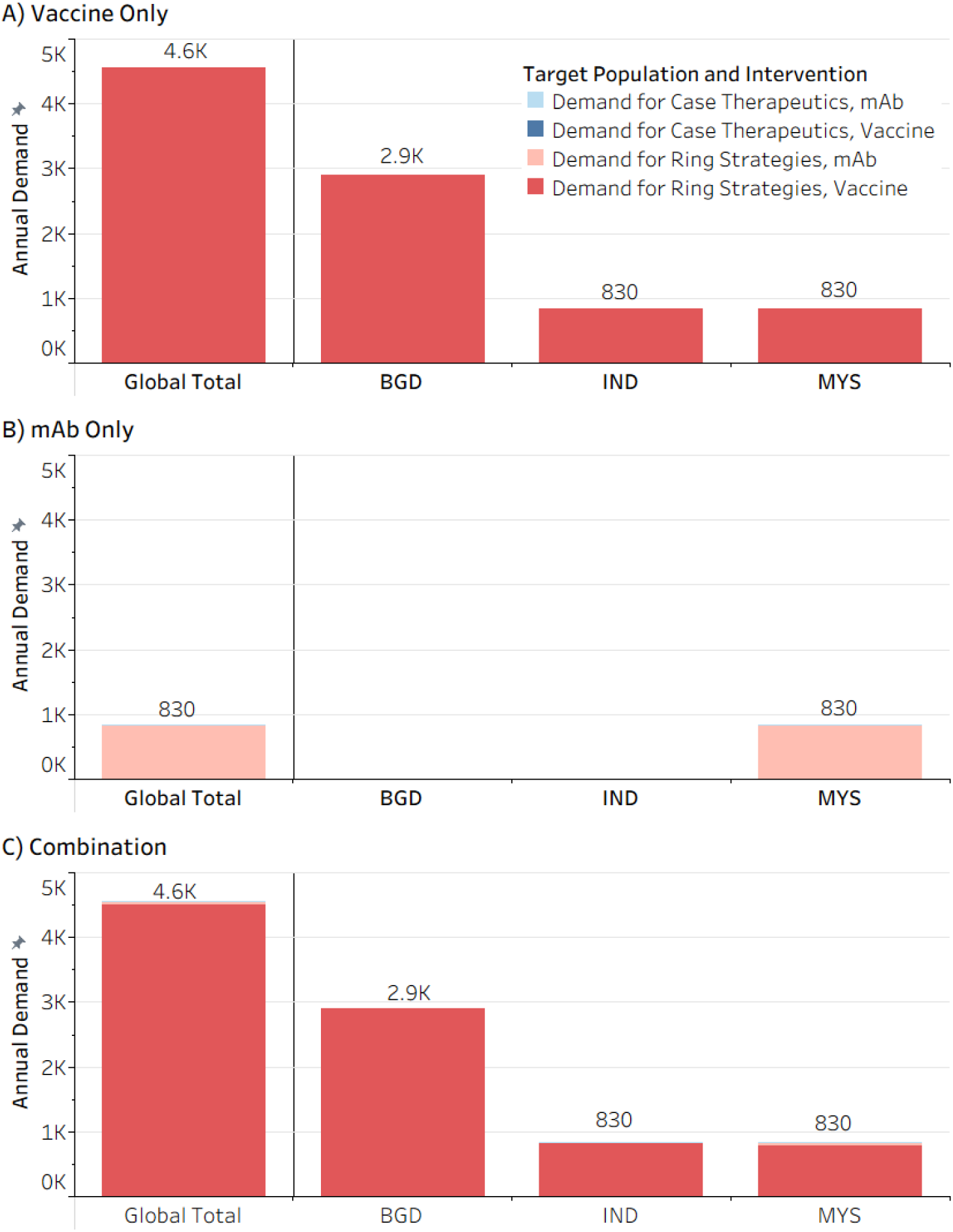
Global annual demand by intervention in response to Expected outbreaks (Scenario 1) in affected countries. Demand is shown assuming the available interventions are vaccine-only, mAb-only, or a combination of the two. Country response is assumed to include ring strategies (including contacts and contacts of contacts) and case therapeutics (for mAbs only). The affected countries include Bangladesh (7 cases), India (2 cases), and Malaysia (2 cases). Note that the global demand total may not exactly match the sum across all countries due to rounding.

##### Country-Level Demand Amplified (Scenario 2) and Extended (Scenario 3) Outbreaks

In active outbreak countries (Bangladesh and India), demand for MCMs increased with outbreak size under the Amplified and Extended outbreak assumptions (Figure 2). In Bangladesh, demand projections reflected stakeholder-reported cost constraints and a preference for established interventions. Total demand ranged from 0 doses (for mAb-only responses to an Amplified outbreak) to 2,100,000 doses in an Extended outbreak. For vaccine-only and combination interventions, reactive ring vaccination was implemented in both outbreak scenarios, with demand ranging from 9,500 to 62,000 doses, respectively. However, in the mAb-only scenario, ring deployment occurred only in the Extended outbreak, consistent with reported hesitancy toward broader mAb use. Proactive vaccination of HCWs (18,000 doses) and rural populations (2,000,000 doses) was introduced for the Extended outbreak only for vaccine-only and combination interventions. Vaccines, assumed to cover everything except case therapeutics, accounted for greater than 99% of total demand in the combination intervention response. For the mAb-only intervention, case therapeutics, ring strategies, and prophylactic campaigns for HCWs and rural populations were introduced only for the Extended outbreak.

**Figure 2.**
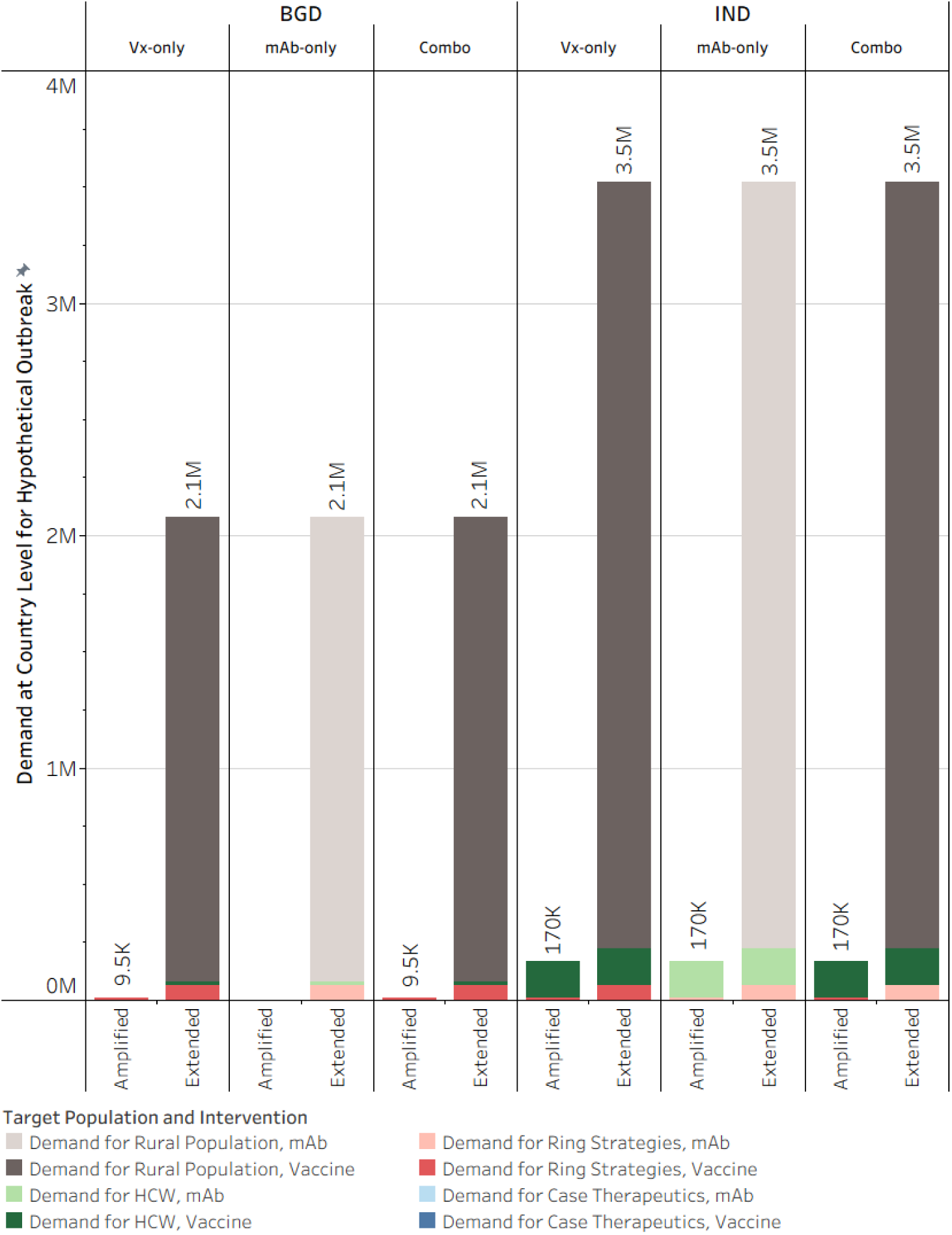
Country-level demand for active outbreak countries in response to Amplified (Scenario 2) and Extended (Scenario 3) outbreaks. The active outbreak countries include Bangladesh (BGD) and India (IND). Demand estimates assume available interventions are vaccine-only, mAb-only, or a combination of the two. The country response is spread across case therapeutics, a reactive ring strategy, and proactive campaigns for HCWs or rural male populations. Note that the country-level total may not sum exactly to the total across all target populations due to rounding.

In India, demand reflected stakeholder preference for vaccines and willingness to implement earlier, more intensive responses. Total demand ranged from 170,000 doses in an Amplified outbreak to 3,500,000 doses in an Extended outbreak. The lowest target population was for mAb therapeutics for infected cases across both outbreak scenarios (30–200 doses). Across both Amplified and Extended outbreaks, vaccines and mAbs were used in ring strategies for vaccine-only and mAb-only interventions, respectively (9,500–62,000 doses). Proactive campaigns for HCWs were also introduced in both intervention scenarios in the Amplified outbreak (160,000 doses). For the combination intervention, mAbs were primarily reserved for case treatment in the Amplified outbreak and then expanded to ring use in the Extended outbreak, representing 0.02%–2% of the total demand. In the Extended outbreak, proactive campaigns across all intervention assumptions expanded to include rural populations (3,300,000 doses).

In past outbreak countries (Malaysia, the Philippines, and Singapore), demand increased in response to larger outbreaks while reflecting KII sentiment around previous outbreak experience, funding constraints, and intervention preferences (Figure 3). In Malaysia, the modeled response reflected stakeholder interest in mAbs for both therapeutic and prophylactic use, as well as support for aggressive responses informed by prior outbreak experience. Total demand ranged from 85,000 to 350,000 doses for Amplified and Extended outbreaks, respectively. Reactive demand from ring strategies ranged from 9,500 to 62,000 doses across Amplified and Extended outbreaks and was applied consistently across vaccine-only and mAb-only strategies. The mAb-only reactive demand also included 30-200 doses for immediate cases. Proactive campaigns for HCWs began in the Amplified outbreak (75,000 doses) and expanded to rural populations in the Extended outbreak (210,000 doses). In the combination intervention, mAbs were allocated to case treatment (30–200 doses) and to ring strategies in both outbreaks (9,500–62,000 doses), while vaccines covered proactive campaigns (75,000-210,000 doses). For this combination intervention, mAbs accounted for approximately 11%–18% of total demand.

**Figure 3.**
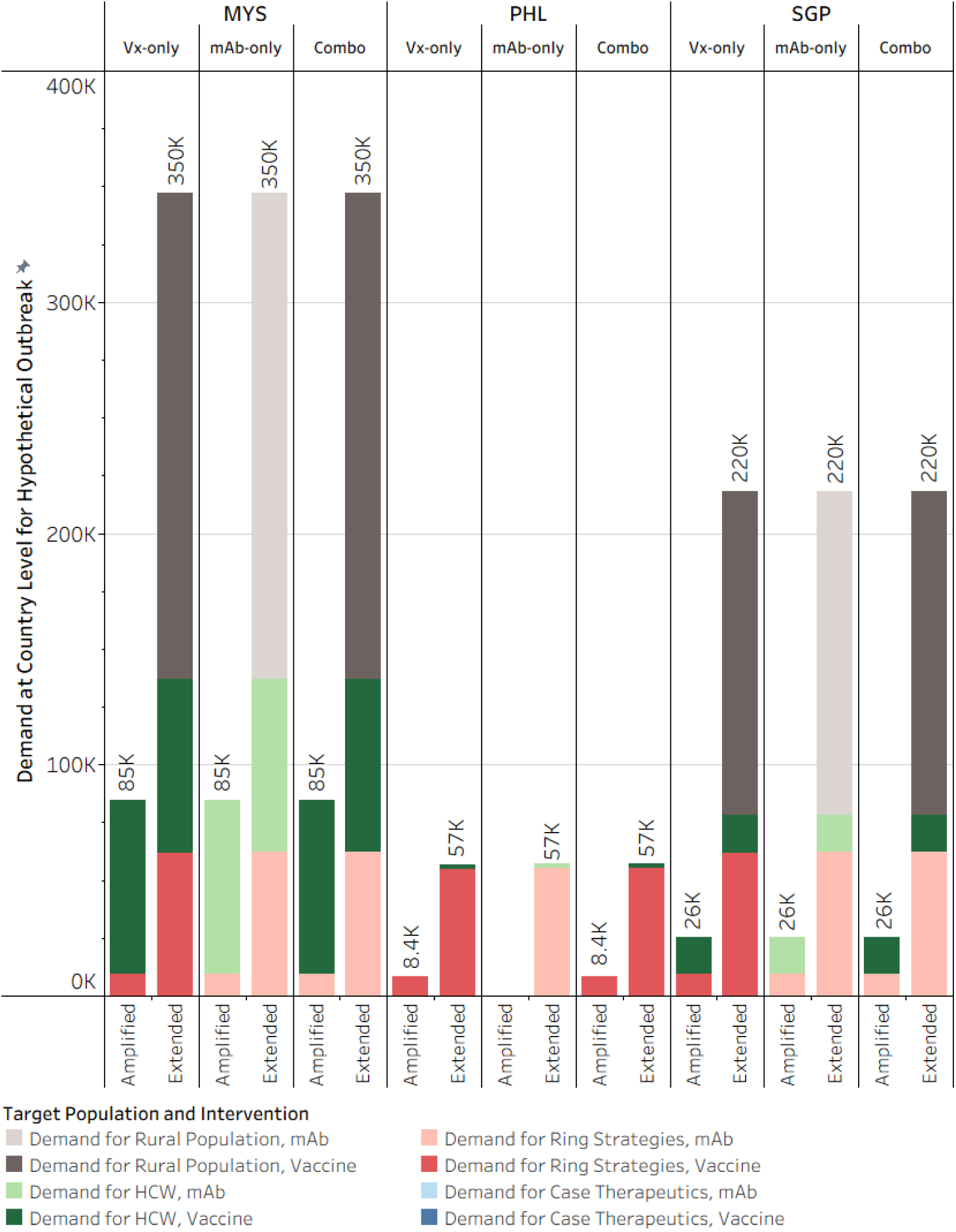
Country-level demand for past outbreak countries in response to Amplified (Scenario 2) and Extended (Scenario 3) outbreaks. The past outbreak countries include Malaysia (MYS), the Philippines (PHL), and Singapore (SGP). Demand estimates assume available interventions are vaccine-only, mAb-only, or a combination of the two. The country response is spread across case therapeutics, a reactive ring strategy, and proactive campaigns for HCWs or rural male populations. Note that the country-level total may not sum exactly to the total across all target populations due to rounding.

In the Philippines, limited surveillance and financing constraints delayed the scale-up of interventions, with total demand ranging from 0 doses (for the mAb-only response to an Amplified outbreak) to 57,000 doses in the Extended outbreak. In vaccine-only scenarios, reactive ring vaccination was introduced in both outbreaks (8,400–55,000 doses). In mAb-only scenarios, case therapeutics (180 doses) and ring vaccination (55,000 doses) were introduced only in the Extended outbreak. Proactive healthcare worker vaccination was also limited to the Extended outbreak (2,000 doses), and rural populations were not targeted. In the combination intervention, mAbs were restricted to case treatment during the Extended outbreak (180 doses), representing less than 1% of total demand, while vaccines covered ring and proactive campaigns.

In Singapore, stakeholders preferred vaccines for large outbreaks and mAbs for limited use. Total demand ranged from 26,000 to 220,000 doses for Amplified and Extended outbreaks, respectively. In vaccine-only scenarios, reactive ring vaccination was implemented in both outbreaks (9,500– 62,000 doses), with proactive healthcare worker vaccination beginning in the Amplified outbreak (16,000 doses) and rural campaigns added in the Extended outbreak (140,000 doses). In mAb-only scenarios, similar reactive and proactive strategies were applied, with mAbs also used for case therapeutics (30–200 doses). In the combination intervention, mAbs were allocated to case treatment and reactive ring strategies, while vaccines supported proactive campaigns. Accordingly, mAbs comprised 29%–38% of total demand in this combination scenario.

In no known outbreak countries (Australia, Cambodia, Indonesia, Thailand, and Vietnam), proactive approaches were only assumed in an Extended outbreak unless KIIs or desk research provided justification for a larger response (Figure 4). In Australia, assumptions were based on desk research showing a strong outbreak response capability. Reactive ring strategies were used in all outbreaks (9,500–62,000 doses), with proactive HCW vaccinations introduced in the Amplified outbreak (110,000 doses) and expanded to rural populations in the Extended outbreak (69,000 doses). For mAb-only interventions, case therapeutics were included in all outbreaks (30–200 doses). In the combination intervention, mAbs were used for case treatment and ring responses across all outbreaks, while vaccines supported proactive campaigns. mAbs accounted for 8%–26% of total demand in the combined scenarios.

**Figure 4.**
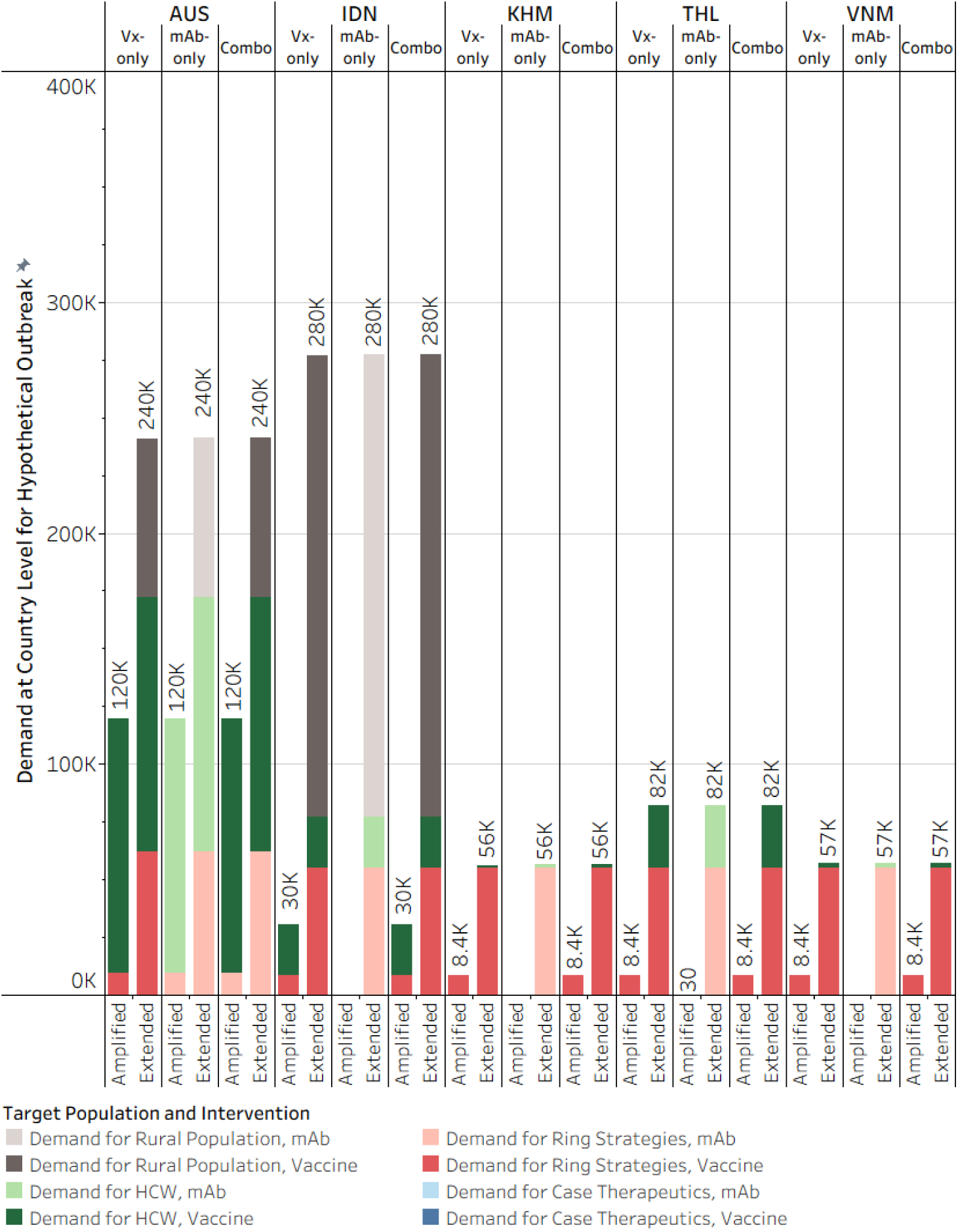
Country-level demand for no known outbreak countries in response to Amplified (Scenario 2) and Extended (Scenario 3) outbreaks. The past outbreak countries include Australia (AUS), Indonesia (IDN), Cambodia (KHM), Thailand (THL), and Vietnam (VNM). Demand estimates assume available interventions are vaccine-only, mAb-only, or a combination of the two. The country response is spread across case therapeutics, a reactive ring strategy, and proactive campaigns for HCWs or rural male populations. Note that the country-level total may not sum exactly to the total across all target populations due to rounding.

In Indonesia, stakeholders expressed concern about regional risk but strongly preferred vaccines. In vaccine-only scenarios, reactive ring vaccination was implemented in all outbreaks (8,400–55,000 doses), with proactive HCW vaccination introduced in the Amplified outbreak (22,000 doses) and rural campaigns in the Extended outbreak (200,000 doses). mAb-only strategies were limited to the Extended outbreak and included case therapeutics (180 doses), ring vaccination (55,000 doses), and both proactive campaigns. In combination scenarios, mAbs were reserved for case treatment, and vaccine deployment followed the same pattern as in vaccine-only scenarios.

In Cambodia, projections reflected low prioritization and price sensitivity determined through desk research. In vaccine-only scenarios, reactive ring vaccination began in the Amplified outbreak (8,400–55,000 doses), with proactive HCW vaccination introduced in the Extended outbreak (1,200 doses). In mAb-only scenarios, case therapeutics (180 doses), ring vaccination (55,000 doses), and HCW campaigns (1,200 doses) were limited to the Extended outbreak. In combination scenarios, mAbs were restricted to case treatment, while vaccines supported reactive and proactive responses.

In Thailand and Vietnam, response and preparedness assumptions were based on KIIs (Thailand) and desk research (Vietnam). In vaccine-only scenarios, reactive ring vaccination began during the Amplified outbreak (8,400–55,000 doses), while proactive HCW vaccination was added in the Extended outbreak (Thailand: 27,000 doses; Vietnam: 1,900 doses). Rural campaigns were not part of any outbreak response. In mAb-only scenarios, case therapeutics were introduced during the Amplified outbreak in Thailand and during the Extended outbreak in Vietnam, with ring and proactive strategies applied only in the Extended outbreak. In combination scenarios, mAbs were used solely for case treatment, while vaccines covered both reactive and proactive efforts.

## 4. Discussion

This analysis estimated the future demand for NiV vaccines and mAbs currently in clinical development by combining country stakeholder interviews with scenario-based epidemiologic modeling. In most countries, expected use was predominantly reactive. Bangladesh and India, which face recurrent outbreaks, along with Malaysia, which has previous outbreak experience, represented all demand under Expected outbreaks. Global annual demand in this scenario ranged from approximately 800-5,000 doses, with response focused mainly on ring strategies due to low case numbers and intermittent outbreaks. In this response, mAbs were mainly used for treating cases, with limited use for prevention.

Under larger Amplified and Extended outbreaks, proactive vaccination of HCWs and rural populations substantially increased total dose requirements. These outbreaks were assumed to occur independently within each country; however, if all countries introduced one-time, proactive campaigns for these target populations, global demand would require 410,000 doses for HCWs and 6,300,000 doses for rural populations. Nonetheless, such strategies were consistently constrained by affordability concerns. Most middle-income countries indicated that large preventive campaigns would be unlikely without external financial support. As a result, under plausible near-term conditions, demand remains episodic, reactive, and geographically concentrated.

### 4.1. Intervention Preferences and Familiarity

No country has a developed strategy for Nipah vaccines or mAbs at this time. However, key informants across all countries generally viewed vaccines as the more practical and cost-effective long-term intervention. mAbs were recognized as clinically valuable but were considered expensive and likely to depend on external donor support in some countries. They were viewed as a complementary tool to vaccines, mainly for therapeutic rather than preventive use, due to their higher cost. For example, representatives from India stated that vaccine development was prioritized over mAbs, which were seen as components of combination therapies rather than standalone solutions. In scenarios favoring vaccines alone or combined, India preferred vaccines over mAbs, which are mainly used to treat infected individuals. Nonetheless, as major MCM manufacturers, Indian companies are likely to produce mAbs internally if vaccine development fails in the mAb-only scenario. Malaysian representatives emphasized the potential of mAbs for dual-use if affordable prices are guaranteed. Singapore reported willingness to pay higher per-dose prices for limited mAb supplies due to their flexibility and smaller target populations, while other countries considered mAbs financially unfeasible for national stockpiles. Our demand forecast reflected this general hesitation toward the widespread use of mAbs, resulting in lower utilization compared to vaccines, even in mAb-only and combined scenarios. Raising awareness of mAb use cases and improving mAb affordability could shift preferences in countries expressing cost constraints, particularly Bangladesh, India, Indonesia, and the Philippines.

Lack of familiarity in country may be partially addressed through clinical trial locations. Country introduction decision-making has been influenced by local applicability and effectiveness, the case for which can be strengthened with domestic clinical trials [37]. For example, as the current location of the Oxford vaccine clinical trial, Bangladesh was expected to be more willing to introduce the vaccine with its limited budget than a mAb product. In India, local manufacturing was perceived as necessary for introduction, as it has strong preferences for locally produced products across its existing vaccine markets. Recently, Serum Institute of India received a license for the development of Oxford’s ChAdOx1 Nipah vaccine, securing country utilization of the vaccine [38]. With this in mind, one avenue to support mAb use could be targeted country selection for clinical trials.

### 4.2. Market viability

MCMs targeting diseases with episodic and low incidence are known to challenge traditional market viability [39]. In the case of Nipah, our findings highlight several challenges. First, only reactive use of vaccines and mAb resulted in annual demand in response to Expected outbreaks, which would be insufficient to sustain continuous manufacturing. Second, the inclusion of proactive interventions in HCW and rural populations increased dose requirements by several orders of magnitude, but these volumes remain insufficient to support cost-efficient continuous manufacturing, particularly for these price-sensitive campaigns. Third, in Amplified or Extended outbreaks, demand can increase to millions of doses in specific countries, posing a challenge to scaling rapidly to meet the expected demand level.

Global institutions and countries will need to align strategies to enable MCM manufacturing readiness for a higher-order outbreak and availability of MCMs for annual cases, while ensuring equitable access for resource-constrained countries. True preparedness includes the willingness to finance readiness for low-probability, high-consequence events. In this sense, NiV resembles other epidemic-prone pathogens like Ebola for which public goods financing, rather than country-level purchasing power, underpins supply [40]. Future work should explicitly examine supply-side perspectives, including manufacturer cost structures, minimum efficient production scales, and acceptable risk-sharing arrangements.

### 4.3. Epidemiologic uncertainty and future risk

Demand estimates in this study were anchored in historical observed epidemiology. However, NiV transmission is influenced by ecological and climatic factors, including habitat disruption for bats, agricultural intensification, and human encroachment. Climate variability and habitat disruption have been associated with spillover dynamics in related zoonotic systems and may plausibly increase the frequency or geographic distribution of NiV outbreaks [41].

If spillover frequency increases modestly, even without changes in transmissibility, reactive demand would rise proportionally. More consequentially, if sustained human-to-human transmission were to occur, even at moderate levels, demand could expand rapidly beyond the levels estimated under expected epidemiology. In Bangladesh, India, and Malaysia, our Amplified and Extended outbreak scenarios provide initial approximations of this expansion relative to baseline demand in the Expected scenario. For ring strategies, each case required roughly 300 additional doses, increasing demand for Amplified outbreaks by roughly 3- to 12-fold and for Extended outbreaks by roughly 20- to 75-fold relative to the baseline. Preventive campaigns, generally targeting HCWs in Amplified outbreaks and adding male rural populations in Extended outbreaks, further increased demand by orders of magnitude relative to the baseline, amounting to hundreds of thousands of doses of demand for HCWs and millions of doses of demand for male rural population.

Another area of uncertainty for these target populations is the location of NiV spillovers. These estimates for preventive campaigns were based on population data for high-risk states or districts with a history of cases or the presence of bat or animal reservoirs. Demand would increase further if surveillance efforts detected a wider geographic spread of cases within the 10 countries included in the analysis. Although speculative, these scenarios are relevant for products in development to inform strategic deployment and underscore the need for preparedness investments to account for uncertainty in future transmission patterns.

### 4.4. Extended populations and willingness to vaccinate

Higher demand estimates were driven primarily by the inclusion of HCWs and male rural populations in proactive campaigns. However, willingness to vaccinate in these groups remains uncertain.

Stakeholder interviews suggested conceptual support for protecting HCWs, particularly in countries with prior outbreak experience, but financing constraints were a consistent barrier. For rural populations, interest was generally conditional on outbreak severity and funding availability. No country reported established plans for routine preventive NiV vaccination.

Accordingly, proactive demand should be interpreted as contingent rather than guaranteed. Without earmarked funding, inclusion of extended populations is unlikely under current fiscal environments. This reinforces the conclusion that baseline, country-financed demand will remain limited in the absence of external support.

### 4.5. Implications for policy and product development

For countries, strengthening surveillance and diagnostic capacity remains essential. Improved detection would both enable earlier, more targeted MCM deployment and reduce the demand uncertainty for reactive and proactive responses. Investments in preparedness infrastructure beyond surveillance and diagnostics will also have carry-over benefits in response to a Nipah outbreak.

For funders and global health institutions, the results support consideration of advance financing mechanisms and globally managed stockpiles. A centralized or regional stockpile, analogous to that used for Ebola vaccines, [42], could ensure equitable and rapid access during outbreaks, aggregate demand across countries, and reduce uncertainty for manufacturers. Without such mechanisms, the low and unpredictable nature of demand may challenge sustained supply. Furthermore, NiV has been designated a priority pathogen by the WHO and CEPI because of its pandemic potential. If a larger outbreak is detected, a globally managed stockpile with advanced financing mechanisms would increase the chances for the timely delivery of MCMs, an essential factor for optimizing their impact [18].

For product developers, the findings indicate that NiV MCMs are likely to be used primarily in reactive, ring-based strategies, with limited to no routine uptake. Product profiles optimized for rapid deployment, ease of storage, and integration into outbreak response systems will be important.

### 4.6. Limitations

Several limitations apply to this work. Stakeholder views may not fully represent national policy positions, and their views may evolve as products advance. Epidemiologic data likely under-detect cases, with surveillance gaps being reported during the most recent outbreak in 2026 [43,44]. This underreporting could potentially lead to an underestimation of demand if detected outbreaks were larger, entire outbreaks were missed, or the geographic spread of high-risk areas in the 10 countries was wider. The case assumptions for each scenario were also based on historical outbreak size, and future outbreaks may exceed the case counts included in our analysis. Demand for these outbreaks could increase compared to our estimates, especially if wide proactive campaigns are adopted. Supply-side perspectives were not systematically incorporated. Finally, scenario assumptions regarding ring strategies and proactive targeting remain subject to debate and could influence dose estimates [45,46].

### 4.7. Conclusions

This study integrated qualitative country perspectives with quantitative scenario modeling to provide a structured, transparent estimate of pre-introduction demand for NiV MCMs. To our knowledge, this study is the first demand forecast for NiV vaccines and mAbs in LMICs and endemic regions. The confirmation of a new fatal Nipah case in Bangladesh and two cases in India in February 2026 [47,48] reinforces the timeliness of this analysis and illustrates how rapidly theoretical preparedness questions become operational realities.

Our central finding is that, under historically observed epidemiology, annual demand is low and episodic, concentrated in reactive ring strategies in a small number of countries. In the expected annual outbreak scenario, demand was limited to approximately 800-5,000 doses globally, rising to the hundreds of thousands or low millions only when proactive vaccination of HCWs and male rural populations was included in response to larger outbreaks. However, these expansions depend on epidemiology, financing, perceived risk, and political prioritization.

The key implication is that NiV MCM markets will be challenging. Demand is most likely contingent on outbreak occurrence, external financing, and stockpile mechanisms. These findings clarify the scale and structure of the market that manufacturers and global funders would need to consider. Addressing this market challenge will require explicit engagement with manufacturers, clearer articulation of acceptable risk-sharing models, and continued assessment of how ecological and climatic change may alter NiV threat trajectories.

## Data Availability

The minimal data set for the underlying assumptions is available in the supporting information.

## Acknowledgments

This work was funded by the Coalition for Epidemic Innovations (CEPI). We acknowledge the CEPI project team (project lead: Arsalan Azmat and project team: Win-Yan Chan and Carolin Vegvari) for their support and continuous feedback throughout the project. We also acknowledge Richard Jarman, Stacey Wooden, Gill Mason, and the wider CEPI Nipah disease team for their valuable comments on the project. Grammarly software (v1.2.279.1925) was used for spelling and grammar checks and for minor syntax and phrasing improvements while editing the manuscript. The authors reviewed and confirmed all changes made by this tool. Grammarly software was limited to language editing in the prepared manuscript only. It was not used in the study design, data collection, data analysis, interpretation of results, or development of conclusions.

## Role of the Funder

This analysis was commissioned by the Coalition for Epidemic Preparedness Innovations (CEPI). The funders advised on country selection but had no other role in study design, data collection, or data analysis. The CEPI project and Nipah disease teams were involved in the decision to publish and provided comments on the manuscript. An earlier version of this analysis was provided to CEPI as an internal report.

## Notes

### Competing Interest Statement

The authors have declared no competing interest.

### Author Declarations

Management Sciences for Health's Scientific Committee determined that the research protocol did not meet the definition of Human Subjects Research and therefore did not require submission to an IRB because the study was designed to collect information about national policies on pandemic preparedness and funding pathways for new vaccines and did not ask participants for information about themselves.

## References

1. Chua KB. Nipah virus: A recently emergent deadly paramyxovirus. Science. 2000;288(5470). doi:10.1126/science.288.5470.1432

2. Luby SP, Gurley ES, Hossain MJ. Transmission of human infection with nipah virus. 2009. doi:10.1086/647951

3. Hossain MJ, Gurley ES, Montgomery JM, Bell M, Carroll DS, Hsu VP, et al. Clinical Presentation of Nipah Virus Infection in Bangladesh. Clin Infect Dis. 2008 Apr 1;46(7):977–84. doi:10.1086/529147

4. Lo MK, Lowe L, Hummel KB, Sazzad HMS, Gurley ES, Hossain MJ, et al. Characterization of Nipah Virus from Outbreaks in Bangladesh, 2008–2010. Emerg Infect Dis. 2012 Feb;18(2):248–55. doi:10.3201/eid1802.111492 PubMed PMID: 22304936; PubMed Central PMCID: PMC3310473.

5. Nikolay B, Salje H, Hossain MJ, Khan AKMD, Sazzad HMS, Rahman M, et al. Transmission of Nipah Virus — 14 Years of Investigations in Bangladesh. N Engl J Med. 2019;380(19). doi:10.1056/nejmoa1805376

6. Arankalle VA, Bandyopadhyay BT, Ramdasi AY, Jadi R, Patil DR, Rahman M, et al. Genomic characterization of nipah virus, West Bengal, India. Emerg Infect Dis. 2011;17(5). doi:10.3201/eid1705.100968

7. Gurley ES, Montgomery JM, Hossain MJ, Bell M, Azad AK, Islam MR, et al. Person-to-person transmission of Nipah virus in a Bangladeshi community. Emerg Infect Dis. 2007;13(7). doi:10.3201/eid1307.061128

8. Islam MS, Sazzad HMS, Satter SM, Sultana S, Hossain MJ, Hasan M, et al. Nipah virus transmission from bats to humans associated with drinking traditional liquor made from date palm sap, Bangladesh, 2011-2014. Emerg Infect Dis. 2016;22(4). doi:10.3201/eid2204.151747

9. World Health Organization. Pathogens prioritization: a scientific framework for epidemic and pandemic research preparedness [Internet]. Geneva; 2024 Jun [cited 2026 May 15]. Available from: https://www.who.int/publications/m/item/pathogens-prioritization-a-scientific-framework-for-epidemic-and-pandemic-research-preparedness

10. Coalition for Epidemic Preparedness Innovations. Nipah virus: The deadly illness without a vaccine. 2021.

11. Coalition for Epidemic Preparedness Innovations. Our portfolio [Internet]. [cited 2025 Dec 11]. Available from: https://cepi.net/our-portfolio

12. Coalition for Epidemic Preparedness Innovations. AI-enhanced self-amplifying mRNA vaccine set to combat one of the deadliest known viruses [Internet]. [cited 2025 Dec 11]. Available from: https://cepi.net/ai-enhanced-self-amplifying-mrna-vaccine-set-combat-one-deadliest-known-viruses

13. Ajelli M, Muyembe JJ, Touré A, Diallo A, Litvinova M, Merler S, et al. Vaccination strategies for Ebola in the democratic republic of Congo: the who-Ebola modeling collaboration. Int J Infect Dis. 2025 Apr;153. doi:10.1016/j.ijid.2025.107779 PubMed PMID: 39805421.

14. Hogan AB, Winskill P, Watson OJ, Walker PGT, Whittaker C, Baguelin M, et al. Within-country age-based prioritisation, global allocation, and public health impact of a vaccine against SARS-CoV-2: A mathematical modelling analysis. Vaccine. 2021;39(22). doi:10.1016/j.vaccine.2021.04.002

15. Moore S, Hill EM, Dyson L, Tildesley MJ, Keeling MJ. Modelling optimal vaccination strategy for SARS-CoV-2 in the UK. PLoS Comput Biol. 2021;17(5). doi:10.1371/journal.pcbi.1008849

16. World Health Organization. Principles and considerations for adding a vaccine to a national immunization program: from decision to implementation and monitoring: From decision to implementation and monitoring [Internet]. 2014 Apr. Available from: https://www.who.int/publications/i/item/9789241506892

17. World Health Organization. WHO target product profile for Nipah virus vaccines [Internet]. 2017 Jun [cited 2026 May 15]. Available from: https://www.who.int/publications/m/item/who-target-product-profile-for-nipah-virus-vaccines

18. Cortes-Azuero O, Vegvari C, Sutcliffe E, Roney E, Scarponi D, Mukandavire C, et al. Modeling optimal deployment strategies for Nipah vaccines and monoclonal antibodies. Vaccine. 2026 Jun 20;84:128688. doi:10.1016/j.vaccine.2026.128688

19. Moradpour J, Shajarizadeh A, Carter J, Chit A, Grootendorst P. The impact of national income and vaccine hesitancy on country-level COVID-19 vaccine uptake. PLOS ONE. 2023 Nov 2;18(11):e0293184. doi:10.1371/journal.pone.0293184

20. Bhatt A, Monk V, Bhatti A, Eiden AL, Hermany L, Hansen N, et al. Identifying factors that can be used to assess a country’s readiness to deploy a new vaccine or improve uptake of an underutilised vaccine: a scoping review [Internet]. 2024 May 1. doi:10.1136/bmjopen-2023-080370

21. World Health Organization. Nipah virus [Internet]. 2026 [cited 2025 Jun 20]. Available from: https://www.who.int/news-room/fact-sheets/detail/nipah-virus

22. Sun YQ, Zhang YY, Liu MC, Chen JJ, Li TT, Liu YN, et al. Mapping the distribution of Nipah virus infections: a geospatial modelling analysis. Lancet Planet Health. 2024 Jul;8(7):e463–75. doi:10.1016/S2542-5196(24)00119-0

23. Katz IT, Essien T, Marinda ET, Gray GE, Bangsberg DR, Martinson NA, et al. Antiretroviral therapy refusal among newly diagnosed HIV-infected adults. AIDS Lond Engl. 2011 Nov 13;25(17):2177–81. doi:10.1097/QAD.0b013e32834b6464 PubMed PMID: 21832935; PubMed Central PMCID: PMC3272300.

24. United Nations Children’s Fund. Immunization Supply Chain Interventions to Enable Coverage and Equity in Urban Poor, Remote Rural and Conflict Settings [Internet]. 2020 [cited 2025 Nov 12]. Available from: https://www.unicef.org/media/96611/file/Immunization%20supply%20chain%20inter ventions.pdf

25. Perera SM, Garbern SC, Mbong EN, Fleming MK, Muhayangabo RF, Ombeni AB, et al. Perceptions toward Ebola vaccination and correlates of vaccine uptake among high-risk community members in North Kivu, Democratic Republic of the Congo. PLOS Glob Public Health. 2024 Jan 18;4(1):e0002566. doi:10.1371/journal.pgph.0002566 PubMed PMID: 38236844; PubMed Central PMCID: PMC10796044.

26. World Health Organization. National health workforce accounts database [Internet]. 2025. Available from: https://apps.who.int/nhwaportal/

27. Ministry of Health and Family Welfare Bangladesh, WHO. Health labour market analysis in Bangladesh 2021. 2021.

28. Bisanzio D, Davis AE, Talbird SE, Van Effelterre T, Metz L, Gaudig M, et al. Targeted preventive vaccination campaigns to reduce Ebola outbreaks: An individual-based modeling study. Vaccine. 2023 Jan 16;41(3):684–93. doi:10.1016/j.vaccine.2022.11.036

29. World Health Organization. Immunization Handbook, Unit 4: Cold chain and logistics management [Internet]. 2017 [cited 2025 Nov 12]. Available from: https://cdn.who.int/media/docs/default-source/searo/india/publications/immunization-handbook-107-198-part2.pdf

30. United Nations. World Population Prospects 2024: Data Sources [Internet]. 2024. Available from: https://population.un.org/wpp/

31. United Nations. World Urbanization Prospects: The 2018 Revision [Internet]. United Nations, Department of Economic and Social Affairs, Population Division; 2025. Available from: https://population.un.org/wup/downloads?tab=Archive

32. Deka MA, Morshed N. Mapping Disease Transmission Risk of Nipah Virus in South and Southeast Asia. Trop Med Infect Dis. 2018 May 30;3(2):57. doi:10.3390/tropicalmed3020057 PubMed PMID: 30274453; PubMed Central PMCID: PMC6073609.

33. World Health Organization. Immunization Data [Internet]. [cited 2026 Mar 8]. Immunization expenditure. Available from: https://immunizationdata.who.int/global/wiise-detail-page

34. Queensland Government. Business Queensland [Internet]. 2025 [cited 2026 May 29]. Nipah virus. Available from: https://www.business.qld.gov.au/industries/farms-fishing-forestry/agriculture/biosecurity/animals/diseases/guide/nipah-virus

35. Lâm S, Dang-Xuan S, Unger F, Meeyam T, Pham-Duc P, Wacharapluesadee S, et al. Operationalizing regional One Health initiatives in Southeast Asia: Ways forward. One Health. 2025 Apr 14;20:101034. doi:10.1016/j.onehlt.2025.101034 PubMed PMID: 40342871; PubMed Central PMCID: PMC12059403.

36. Pham-Thanh L, Nhu TV, Nguyen TV, Tran KV, Nguyen KC, Nguyen HT, et al. Zoonotic pathogens and diseases detected in Vietnam, 2020–2021. One Health. 2022 Jun 1;14:100398. doi:10.1016/j.onehlt.2022.100398

37. PATH. Approaching Vaccination from End to End - Lesson 4: Decide with data [Internet]. [cited 2025 Nov 16]. Available from: https://www.path.org/edging-out-je/approaching-vaccination-from-end-to-end/lesson-4-decide-with-data/

38. Coalition for Epidemic Preparedness Innovations. Establishing the world’s largest Nipah virus vaccine reserve [Internet]. 2025 [cited 2025 Nov 17]. Available from: https://cepi.net/establishing-worlds-largest-nipah-virus-vaccine-reserve

39. Bloom DE, Cadarette D, Tortorice DL. An Ounce of Prevention: Our approach to vaccine finance is ill-suited to addressing epidemic risk. Finance C Development [Internet]. 2020 Sep [cited 2026 Jun 8]. Available from: https://www.imf.org/en/publications/fandd/issues/2020/09/vaccine-finance-epidemics-and-prevention-bloom

40. UNICEF. Emergency stockpile availability report - Ebola vaccine | UNICEF Supply Division [Internet]. 2026 [cited 2026 Mar 17]. Available from: https://www.unicef.org/supply/documents/emergency-stockpile-availability-report-ebola-vaccine

41. Khan S, Akbar SMF, Mahtab MA, Uddin MdN, Rashid MdM, Yahiro T, et al. Twenty-five years of Nipah outbreaks in Southeast Asia: A persistent threat to global health. IJID Reg. 2024 Dec 1;13:100434. doi:10.1016/j.ijregi.2024.100434

42. World Health Organization. Ebola vaccine stockpiles [Internet]. [cited 2026 Jun 2]. Available from: https://www.who.int/groups/icg/ebola-virus-disease/ebola-stockpiles

43. Hegde ST, Salje H, Sazzad HMS, Hossain MJ, Rahman M, Daszak P, et al. Using healthcare-seeking behaviour to estimate the number of Nipah outbreaks missed by hospital-based surveillance in Bangladesh. Int J Epidemiol. 2019 Aug 1;48(4):1219–27. doi:10.1093/ije/dyz057

44. Chakraborty C, Bhattacharya M, George Priya Doss C, Nandi SS. Re-emergence of Nipah Virus in Eastern India: Urgent need for enhanced surveillance. New Microbes New Infect. 2026 Feb 3;70:101718. doi:10.1016/j.nmni.2026.101718 PubMed PMID: 41732519; PubMed Central PMCID: PMC12925422.

45. Strategic Advisory Group of Experts (SAGE) on Immunization. Interim Recommendations on Vaccination against Ebola Virus Disease (EVD) [Internet]. 2019 [cited 2026 Jan 30]. Available from: https://cdn.who.int/media/docs/default-source/immunization/ebola/interim-ebola-recommendations-may-2019.pdf

46. Muyembe JJ, Pan H, Peto R, Diallo A, Touré A, Mbala-Kingebene P, et al. Ebola Outbreak Response in the DRC with rVSV-ZEBOV-GP Ring Vaccination. N Engl J Med. 2024 Dec 18;391(24):2327–36. doi:10.1056/NEJMoa1904387

47. Nipah virus infection - Bangladesh [Internet]. 2026 [cited 2026 Feb 9]. Available from: https://www.who.int/emergencies/disease-outbreak-news/item/2026-DON594

48. Nipah virus disease - India [Internet]. 2026 [cited 2026 Feb 9]. Available from: https://www.who.int/emergencies/disease-outbreak-news/item/2026-DON593

